# Evaluation of the efficacy of *Lactobacillus*-containing feminine hygiene products on vaginal microbiome and genitourinary symptoms in pre- and postmenopausal women: A pilot randomized controlled trial

**DOI:** 10.1101/2022.06.08.22276136

**Authors:** Remi Yoshikata, Michiko Yamaguchi, Yuri Mase, Ayano Tatsuyuki, Khin Z. Myint, Hiroaki Ohta

## Abstract

**Objective:** As estrogen level decreases with aging, the vaginal mucosa gets thinner, and collagen amount decreases. In addition, the population of *Lactobacillus* in the vagina declines, increasing the risk of atrophic vaginitis, bacterial vaginosis, and genitourinary symptoms in the postmenopausal women. In this study, we evaluated the effects of *Lactobacillus*-containing feminine hygiene products on vaginal microbiome and genitourinary symptoms in pre- and postmenopausal women.

**Methods:** This was a pilot randomized controlled trial in 35 premenopausal and 35 postmenopausal healthy women. For 4 weeks, treatment 1 group (14 premenopausal and 16 postmenopausal women) used the Lactobacillus-containing feminine soap and cream, and treatment 2 group (15 premenopausal and 14 postmenopausal women) used Lactobacillus-containing feminine gel in addition to soap and cream. The remaining 6 premenopausal and 5 postmenopausal women served as controls without using any products. We then compared the changes in the vaginal microbiota, genitourinary symptoms, and other related biomarkers after completion of treatment.

**Results:** Vaginal pH and pathogenic flora were reduced in both treatment groups compared to control group. It was more significant in the treatment 2 group of postmenopausal women. Genitourinary symptoms significantly improved in 60% of premenopausal women in treatment 1 group and 81.3% of postmenopausal women in treatment 2 group, compared to control group (0%). Overactive bladder symptom scores were significantly improved after using the products in the postmenopausal women suspected of having overactive bladder.

**Conclusion:** The use of Lactobacillus-containing feminine products was associated with improved vaginal ecosystem and urogenital health compared to control group, especially in those women using feminine gel.

## Introduction

*Lactobacillus* species are highly ubiquitous bacteria, belonging to the group of lactic acid bacteria (LAB). They are widely found in fermented food as well as in humans and animals. They produce lactic acids, thus lower pH and inhibit the growth of bacteria. Therefore, *Lactobacillus* species, through their natural protective actions, maintain the ideal vaginal environment and vaginal health.^1^ In female vagina, there are five community types or community state types (CSTs) of bacteria.^2^ Among them four are *Lactobacillus* dominant types with low microbial diversity: *Lactobacillus crispatus* (CST I), *Lactobacillus gasseri* (CST II), *Lactobacillus iners* (CST III) and *Lactobacillus jensenii* (CST V). The ranking of Lactobacillus was in the order of CST I, II, V, and III. The better the rank was, the lower the risk of sexually transmitted infections and viral infections, such as HIV, HSV, HPV, gonorrhea, chlamydia, and trichomonas. ^3^ The last one is the CST IV or diversity type. This type has a greater diversity of bacterial taxa with low proportions of typical *Lactobacillus* species. CST IV can be further clustered into CST IV-A and CST IV-B, with respect to different compositions of bacterial species. ^4^ Diversity type is associated with an increased risk for various gynecological problems. ^5-7^

The prevalence of CSTs varies across different ethnic groups or geographical locations. ^2^ In addition to this spatial heterogeneity, it possesses temporary heterogeneity as it can change in the same individual across time, in response to the level of circulating estrogen. ^8^ As blood estrogen declines with age, *Lactobacillus* population is dramatically reduced. In addition, estrogen decline is associated with the loss of collagen and elastin, thinning of the vaginal epithelium and reduced elasticity.^9^ These changes also occur in the urinary tract because it has the same embryonic origin as the genital tract. As a result, there are increased risks of bacterial vaginosis and genitourinary symptoms of menopause (GSM).^10^

GSM results from anatomical, histological and functional changes of the female genitourinary system due to reduced estrogen levels and ageing. GSM includes genital symptoms (dryness, burning, itching, irritation, bleeding), sexual symptoms (dyspareunia and other sexual dysfunctions) and urinary symptoms (dysuria, frequency, urgency, recurrent urinary infections). ^11^Treatments for GSM are usually effective if started as early as possible at the onset of the signs and symptoms. Treatment options include both hormonal and non-hormonal therapies, as well as physical therapies. However, most of the women with GSM remained unrecognized and untreated. ^12, 13^

Moderate to severe genitourinary symptoms might need hormonal therapies such as local estrogen treatment. However, probiotics therapies have emerged in recent days as an alternative method to protect urogenital health by improving the microbiological ecology in the genitourinary tract. Lactobacillus are empirically selected for such use. One pilot study showed that topical application of a *Lactobacillus*-ointment can increase the colonization of Lactobacillus in the external and internal genital areas of postmenopausal women up to 10 days after cessation of treatment. ^14^ Several studies have also reported that Lactobacillus-containing probiotic hygienic products were effective for prevention and treatment of urogenital infection and maintenance of urogenital health. ^15^ *Lactobacillus plantarum* and *Lactobacillus rhamnosus* are the most documented, well-studied probiotic strains for use in healthcare products. ^16, 17^ They were also normal commensals found in the healthy vaginal ecosystem ^18^ These human-derived strains are also promising probiotics to enhance urogenital health and shown to have high adhesion ability to vaginal and cervical epithelial cells.^19^

In this pilot study, we aimed to compare the changes in vaginal microbiota and microbial diversity in the pre and postmenopausal women before and after using these Lactobacillus-containing feminine hygiene products. Then we evaluated their efficacy on genitourinary and overactive bladder symptoms.

## Methods

### Study population

This is a pilot randomized controlled trial comprising 70 healthy Japanese women. The recruitment process started in March 2021, using pamphlets and posters targeting the female employees at the Resorttrust, Inc., their relatives and friends. Within one month of recruitment, seventy-three women registered for participation in the study. Among them, 70 were selected based on the inclusion and exclusion criteria as shown in table 1 by the end of March, 2021. Half of them were premenopausal women, and half were postmenopausal women. Menopause was defined as no menstruation at least 12 months since the last menstrual period, follicular stimulating hormone level of 25mIU/mL and above and estradiol level less than 20 pg/ml.

**Table 1.** Inclusion and exclusion criteria of the participants

| Inclusion criteria | Exclusion criteria |
| --- | --- |
| -Pre-menopausal women between 20-49 years of age<br>-Postmenopausal women between 50-75 years of age<br>-Who give consent on the research | 1. Those under treatment for urinary tract infection<br>2. Those under treatment for urinary stone, hydronephrosis, renal tumor, etc.<br>3. Those who have taken antibiotics or steroids 2 weeks before the study or during the study<br>4. Those who have skin lesions on skin and mucosa of external genitalia or gynecological diseases such as vaginitis (except from asymptomatic atrophic vaginitis)<br>5. Those considered ineligible by the investigators |

### Ethical consideration

The Institutional Review Board of Medical Corporation Shinkokai approved the entire research protocol. We explained the details of the study to these women, and obtained their written informed consent. This study was registered with the University Hospital Medical Information Network (UMIN) Clinical Trial Registry (trial registration number: UMIN000043944). The full trial protocol can be accessed in the supplementary information.

### Grouping and treatment details

We listed the premenopausal and postmenopausal women in the chronological order of their registration, and assigned each group into three groups (control, treatment 1 and treatment2) by simple randomization. Control group (6 premenopausal women and 5 postmenopausal women) used no feminine care products. Treatment 1 group (14 premenopausal women and 16 postmenopausal women) used the Lactobacillus-containing feminine foaming wash and cream. Treatment 2 group (15 premenopausal women and 14 postmenopausal women) received Lactobacillus-containing feminine gel in addition to treatment 1. The participants underwent baseline assessments at the Hamasite Clinic, Minato-ku, Tokyo Prefecture, during April and May, 2021. One premenopausal from treatment 1 group dropped out as vaginal assessments were not possible due to menstruation. Therefore, she was substituted with a new participant. Treatment 1 and 2 groups started using the feminine care products on the day after completion of baseline assessments. Treatment 1 group used two pumps of foaming wash (Product name: Delicate Softwash®) while taking bath and gently applied feminine cream (Product name: Delicate Softgelcream®) around the labia majora covering about 2 cm in diameter after bath once a day. Treatment 2 group used vaginal gel (Product name: Inner gel®) with an applicator once in every three days in addition to foaming wash and cream once a day. These feminine products contained *Lactobacillus rhamnosus* vitaP1 and *Lactobacillus plantarum* KCTC3108. All the products were produced by Hanamisui Co., Ltd. and distributed by Advanced Medical Care, Co. Ltd., both of which are based in Tokyo, Japan. The intervention groups reported no adverse or allergic reactions while using or after using these products. They had follow-up assessments after 4-week trial period. Figure 1 showed the CONSORT flow diagram for this pilot randomized controlled trial. The doctors and nurses at Hamasite Medical Clinic generated the random allocation sequence, enrolled participants, and assigned participants to interventions. They also explained, distributed and monitored the use of feminine care products in the intervention groups.

**Figure 1.**
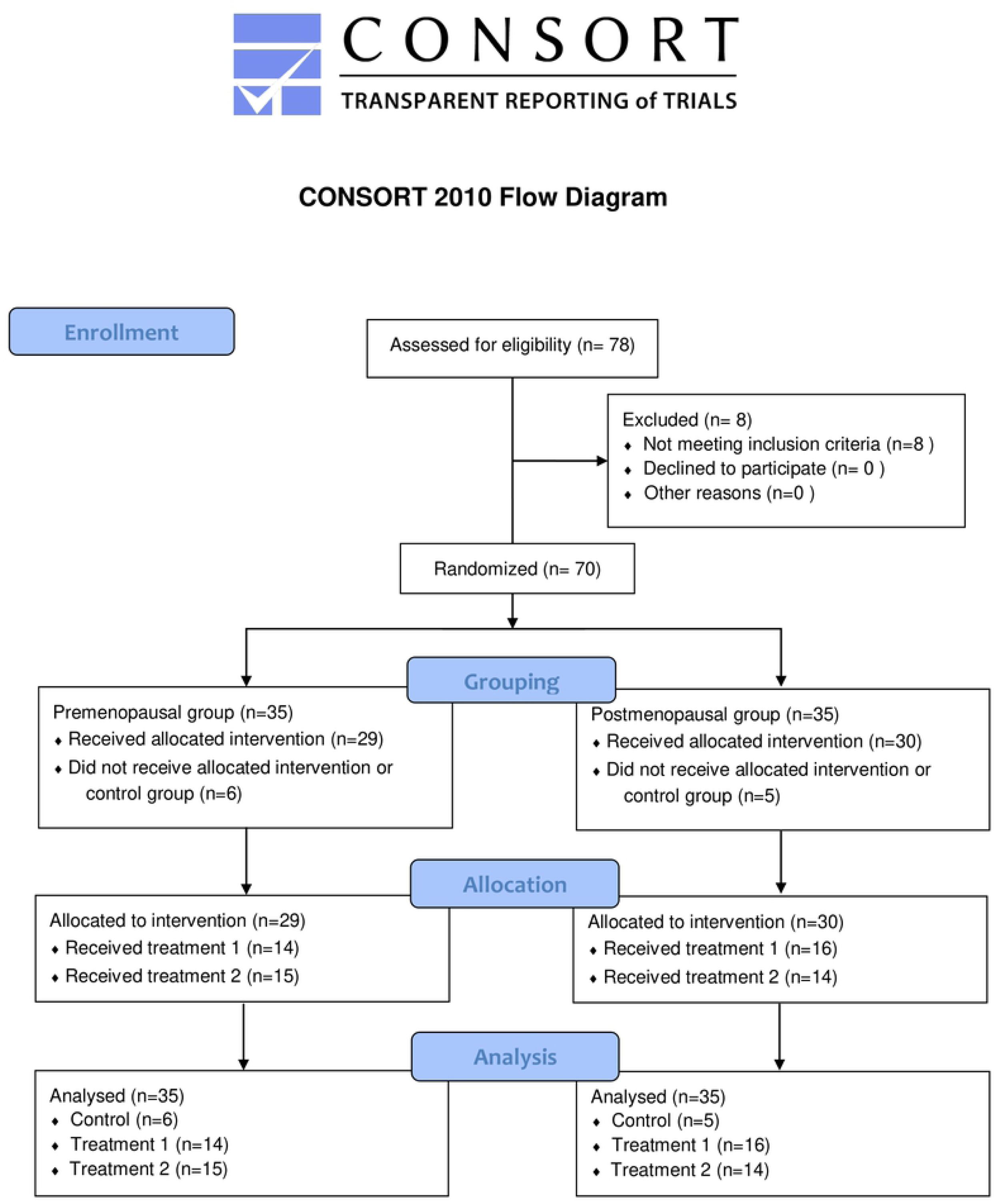

### Measurements

Female sex hormones such as estradiol and follicular stimulating hormone levels were measured from the blood specimens of all participants, using Chemiluminescence ImmunoAssay (CLIA) Test. As there can be physiological fluctuations in women of reproductive age, we collected blood during the ovulation phase in premenopausal women. Gynecologists, who are involved as coinvestigators of this study, took the required samples and pH measurements from the vagina. Vaginal pH was determined by using pH test strips (pH-Fix 3.6–6.1, Macherey-Nagel, Düren, Germany). The measurement of urinary equol was conducted as described by Yoshikata et al.^20^ Women with a urinary equol level higher than 1.0 μM were defined as equol producers. ^21,22^

### Microbiome tests

The experienced gynecologists took the vaginal specimens for microbial analysis. The collected vaginal specimens were put into the container with lysis buffer (4M Guanidine thiocyanate, 40 mM EDTA, 100 mM Tris-HCl pH 9.0) and stored at room temperature. After being transported to the laboratory, they were kept refrigerated and processed within two weeks from the date of collection. The bacterial cell walls were disrupted by a bead beater using zirconia beads to extract or isolate bacterial DNA. The extracted DNA was then purified using an automatic extraction device (Maxwell, Promega). After that approximately 300 base pairs in the V1 and V2 regions of the 16S rRNA fragments were amplified with the V1-V2 region specific primer set 27F/338 R using a thermal cycler (Veriti, Thermo Fisher Scientific, MA, USA). We used a mixture of modified 27F primer to optimize the amplification. The amplified products were then purified and quantified using a real-time PCR device (StepOne™, Thermo Fisher Scientific, MA, USA). A library was prepared using the quantified PCR product, and emulsion PCR was performed using special beads by the OneTouch™ system (Thermo Fisher Scientific, MA, USA). After purifying the special beads, they were loaded into an electronic chip (Ion 318™ Chip) and sequenced using a next-generation sequencer (Ion PGM, Thermo Fisher Scientific, MA, USA). The phylum, class, order, family, genus, and species levels were identified, and the proportion of each species was calculated from the obtained sequences. The details of the protocol were described in previous studies. ^23, 24^ The test results included microbial diversity and distributions of bacterial groups at the phylum, class, order, family, genus, and species levels. Data analysis is performed with Ion Reporter Software. MicroSEQ™ ID 16S rRNA reference database and Greengenes database identified the bacteria down to genus or species level. The alpha diversity scores of the Shannon diversity index were used to describe microbial diversity.

### Questionnaires

All the 70 participants completed self-administered questionnaires regarding their genitourinary symptoms, menopausal symptoms, and overactive bladder symptoms before and after 4 weeks of intervention. Menopausal symptoms were assessed using the simplified menopausal index scale developed by Koyama in 1996. ^25^ It was a commonly used scale in the clinical setting and menopause research in Japan^,26^ which comprises of 4 vasomotor symptoms (facial flushing, sweating, feeling cold, breathlessness and palpitations), 4 psychological symptoms (insomnia, irritability, depression, headaches, dizziness and nausea), and 2 somatic symptoms (tired easily, stiff shoulders and joint pain). These symptoms were evaluated in 4 categories (severe, moderate, mild and never). Some items are given more weight in scoring, and total scores range from 0 to 100, and higher scores indicate more severe symptoms. Also, scores ranging from 0 - 25, 26 - 50, 51

-65, 66 -80 and 81-100 were used to rate the degree of severity and plan for further management. Overactive bladder symptoms were evaluated by using the overactive bladder symptom scores (OABSS).^27, 28^ It assessed the severity of urinary symptoms in scores such as daytime frequency (0 to 2), night-time frequency (0 to 3), urgency (0 to 5), and urgency incontinence (0 to 5). The total score ranges from 0 to 15, and higher scores indicate more severe symptoms (≥ 5, mild; 6–11, moderate; ≥ 12, severe). Supplementary data file 1 showed the details of these self-administered questionnaires.

### Statistical analyses

We estimated the sample size for this study from an 8-week preliminary study on 12 premenopausal and 9 postmenopausal women before and after using feminine care products. We first calculated the effected size for the changes in vaginal microbial diversity as the primary outcome measure. We used that effect size (f=0.53), with the assumption of a two-sided 5% significance level and power of 80%, and calculated that a minimum of 52 women in total was necessary for the comparison (baseline versus week 12) of six groups using a two-way repeated-measures ANOVA (analysis of variance) test. Considering 25% dropout, we recruited 70 people. The sample size calculations, data analysis and figure generation were performed using R software (R 4.1.0, R Core Team, 2021) and Microsoft Excel (Microsoft Corporation, 2018). The distribution of normality was assessed with the Kolmogorov–Smirnov test, box plots, and histograms. Categorical variables were expressed as numbers (N) and percentages (%), continuous variables were expressed as means with standard deviations, and scores were expressed as medians with interquartile ranges. Comparisons between groups were performed using the chi-squared test and Fisher’s exact test for proportions, student’s t-test for continuous variables, and Mann-Whitney U test for scores. The changes in pre-versus post-intervention values in each group were compared using paired t-test for continuous variables, and Wilcoxon Rank sum test for scores. Differences among control and treatment groups were compared using one-way ANOVA test for continuous variables and Kruskal-Wallis test for scores. All tests were two-sided, and statistical significance was set to p < 0.05.

## Results

Table 2 showed the basic characteristics of the participants in our study cohort. Postmenopausal women had higher urinary concentration of equol but lower proportion of equol producers although these findings were not statistically significant. Nevertheless, age-related changes, such as sex hormone levels and vaginal pH (4.37 ± 0.58 versus 5.89 ± 0.45, p<0.0001), were significantly different between the two groups. We also found significant differences in the *Lactobacillus* composition (72% ± 36.84 versus 10% ± 25.11, p<0.0001) as well as vaginal microbial diversity (0.77 ± 0.73 versus 1.92 ± 1.02, p<0.0001) between the two groups of women. Both the control and treatment groups had similar results in these parameters. Baseline simplified menopausal index scores showed no significant differences in any group of women. About 60% of the premenopausal and 50% of postmenopausal women scored less than 25. Another 30% of premenopausal women and 40% of postmenopausal women scored between 26 and 50. Therefore, almost all the women in each group had mild menopausal symptoms without requiring special treatment. Baseline overactive bladder symptom scores (OABSS) were similar between the intervention and control groups. However, postmenopausal women had significantly higher scores compared to the premenopausal women (0.86 versus 2.97, p<.0001). More than 90% of premenopausal women scores less than 3, which is considered having no overactive bladder. However, about 34% of postmenopausal women scored 3 and above, suspected as having overactive bladder (p<0.001).

**Table 2.**
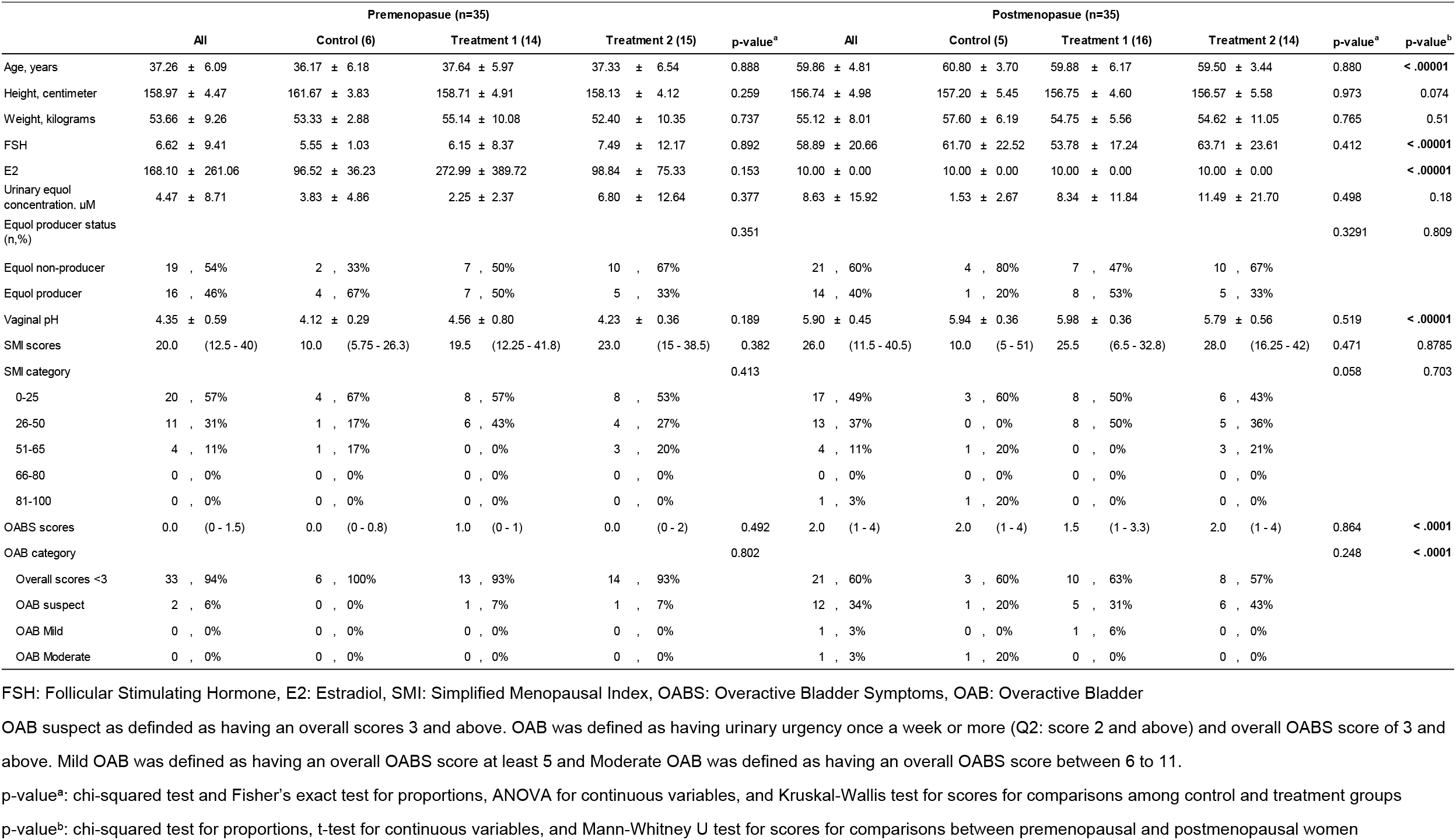
Summary of characteristics of the participants in the study cohort.

Figure 2 showed genitourinary symptoms reported in each group of women and their effects on their daily lives. Almost 60% of women in each group (20 premenopausal women and 22 postmenopausal women) reported that they had at least one genitourinary symptom. In premenopausal women, the most common symptoms were bad odor, itchiness, and increased discharge. Postmenopausal women reported more urinary symptoms such as urinary incontinence, frequency, and dyspareunia. While 70% of premenopausal women rated these effects as mild, 32% of postmenopausal women rated them as moderate, and 18% of postmenopausal women rated these effects as severe.

**Figure. 2.**
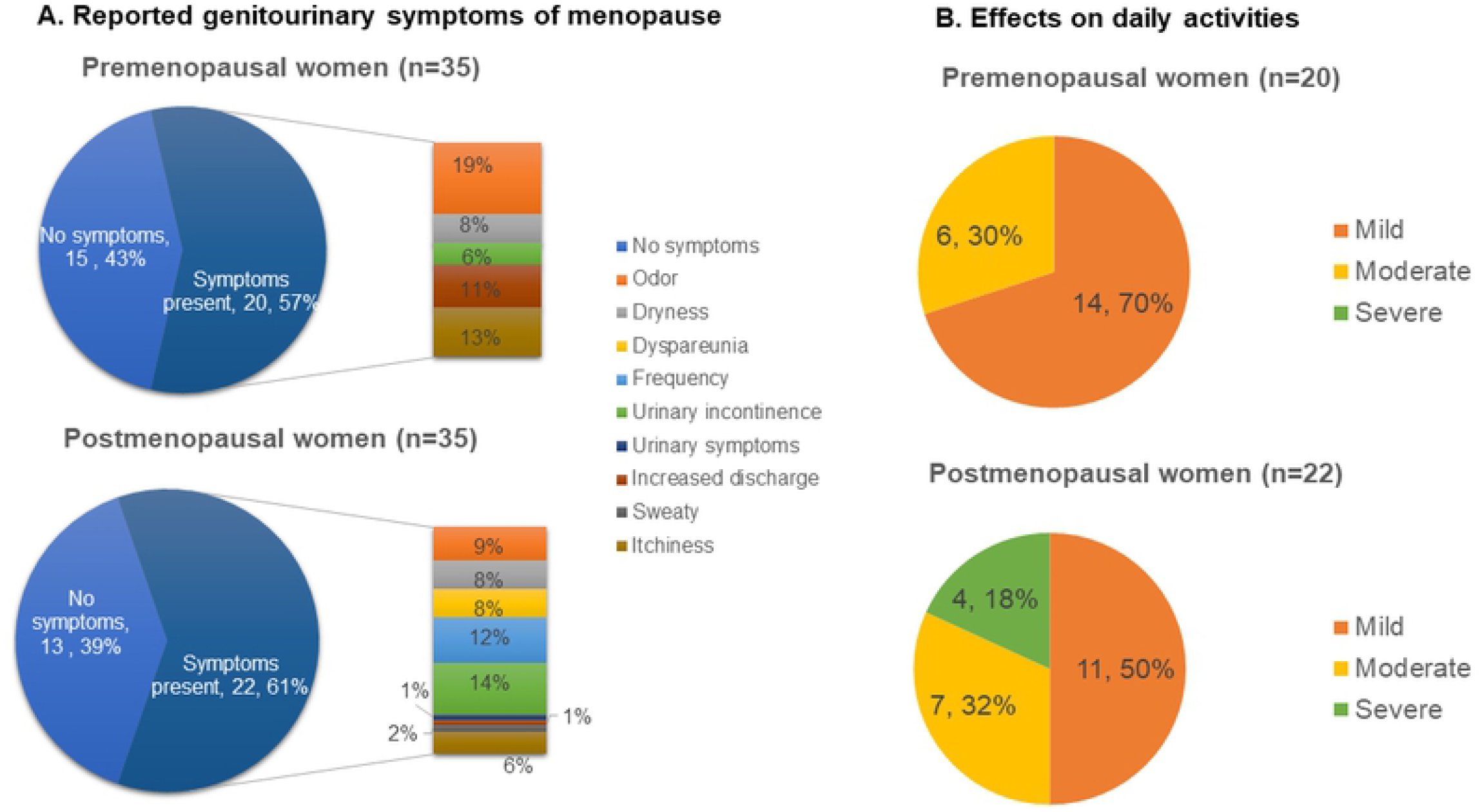
Reported genitourinary symptoms of menopause and their effects on daily activities. Genitourinary symptoms were more varied and stronger in postmenopausal women.

Therefore, postmenopausal women had stronger genitourinary symptoms.

We also found significant differences in the *Lactobacillus* composition (72% ± 36.84 versus 10% ± 25.11, p<0.0001) as well as vaginal microbial diversity (0.77 ± 0.73 versus 1.92 ± 1.02, p<0.0001) between the two groups of women (Table 3). Both the control and treatment groups had similar results in these parameters.

**Table 3.** Baseline vaginal microbiome compositions of the participants in the study cohort

|  | Premenopasue (n=35) |  |  |  |  | Postmenopasue (n=35) |  |  |  |  |  |
| --- | --- | --- | --- | --- | --- | --- | --- | --- | --- | --- | --- |
|  | All | Control (6) | Treatment 1 (14) | Treatment 2 (15) | p-value<sup>a</sup> | All | Control (5) | Treatment 1 (16) | Treatment 2 (14) | p-value<sup>a</sup> | p-value<sup>b</sup> |
| Microbial diversity | 0.77 ± 0.72 | 0.69 ± 0.89 | 0.81 ± 0.82 | 0.76 ± 0.57 | 0.949 | 1.92 ± 1.01 | 2.11 ± 1.15 | 2.03 ± 1.10 | 1.72 ± 0.88 | 0.647 | < .00001 |
| Total *Lactobacillus* , % | 71.98 ± 36.84 | 87.77 ± 19.30 | 64.23 ± 43.89 | 72.90 ± 34.73 | 0.433 | 10.08 ± 25.11 | 6.67 ± 10.07 | 7.36 ± 22.81 | 14.41 ± 31.37 | 0.718 | < .00001 |
| *L. crispatus* (CST I) | 31.71 ± 43.24 | 35.98 ± 48.31 | 29.25 ± 41.94 | 32.29 ± 45.38 | 0.595 | 1.10 ± 6.39 | 7.57 ± 16.92 | 0.04 ± 0.16 | 0.01 ± 0.03 | 0.118 | 0.001 |
| *L. gasseri* (CST II) | 5.02 ± 14.37 | 5.41 ± 13.25 | 1.02 ± 3.83 | 8.60 ± 19.88 | 0.551 | 1.59 ± 7.66 | 0.00 ± 0.00 | 3.48 ± 11.22 | 0.00 ± 0.00 |  | 0.033 |
| *L. iners* (CST III) | 34.54 ± 41.70 | 46.30 ± 51.08 | 39.80 ± 43.80 | 24.93 ± 36.37 | 0.201 | 4.80 ± 19.43 | 0.36 ± 0.50 | 0.07 ± 0.24 | 11.80 ± 29.98 | 0.296 | < .001 |
| *L. jensenii* (CST V) | 0.00 ± 1.57 | 0.00 ± 0.82 | 0.00 ± 1.22 | 0.00 ± 1.92 |  | 0.00 ± 0.02 | 0.00 ± 0.00 | 0.00 ± 0.00 | 0.01 ± 0.03 |  |  |
| Pathogenic bacteria (CST IV) | 14.89 ± 26.27 | 4.48 ± 10.94 | 19.55 ± 32.21 | 14.71 ± 24.54 | 0.515 | 0.77 ± 4.07 | 0.00 ± 0.00 | 0.18 ± 0.71 | 1.73 ± 6.41 | 0.518 | 0.34 |
| Others | 8.36 ± 17.37 | 7.31 ± 16.92 | 5.66 ± 10.24 | 11.29 ± 22.71 | 0.688 | 63.14 ± 34.39 | 55.38 ± 40.70 | 69.98 ± 30.37 | 58.10 ± 37.57 | 0.566 | < .00001 |

p-value<sup>a</sup>: ANOVA for continuous variables for comparisons among control and treatment groups
p-value<sup>b</sup>: t-test for comparisons between premenopausal and postmenopausal women

### Changes after four weeks

Figure 3A and 3B showed the landscape of distribution of pathogenic flora, *Lactobacillus* species and other bacterial communities in premenopausal and postmenopausal women. In premenopausal women, *Lactobacillus* population increased. In general, pathogenic flora population decreased in both groups. Figure 4 showed that vaginal pH and pathogenic flora proportions significantly reduced in postmenopausal women in treatment 2 intervention group (5.79 versus 5.65, p<0.05 and 27.491 versus 4.083, p<0.05, respectively). However, microbial diversity showed a significant increase in treatment 1 group of postmenopausal women (2.03 versus 2.73, p<0.05). One-way ANOVA test showed that pH reduction in treatment 2 postmenopausal group was significant when compared to the postmenopausal control group. Apart from that, no significant differences was observed in vaginal pH, microbial diversity, and proportions of vaginal pathogenic flora in comparison with the control groups.

**Figure. 3.**
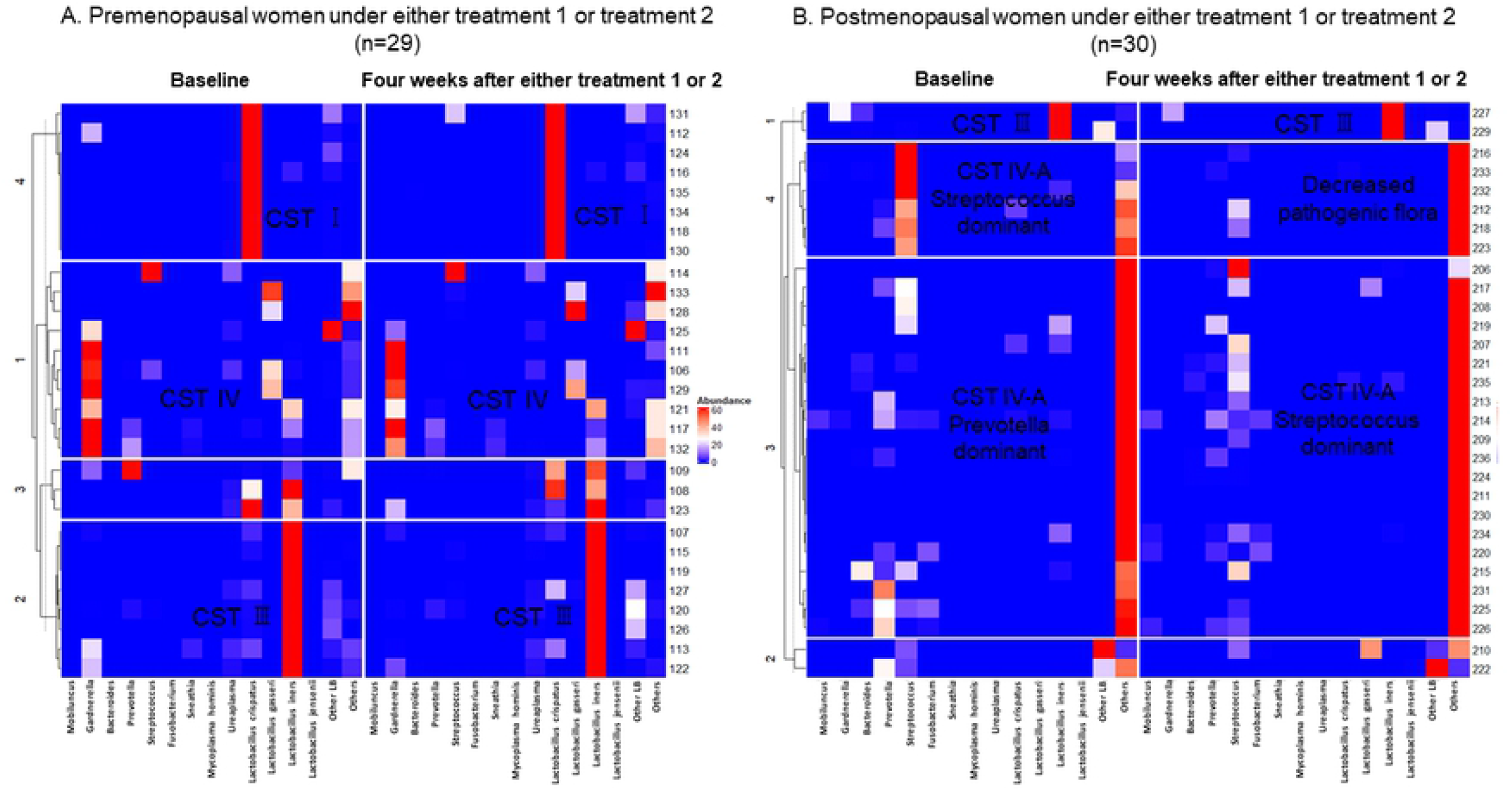
Distribution of pathologic flora and Lactobacillus species in the vagina at baselineand four weeks after intervention. In premenopausal women, Lactobacillus population appeared to be increased, and pathogenic flora decreased in postmenopausal women. In general, pathogenic flora population decreased in both groups.

**Figure. 4.**
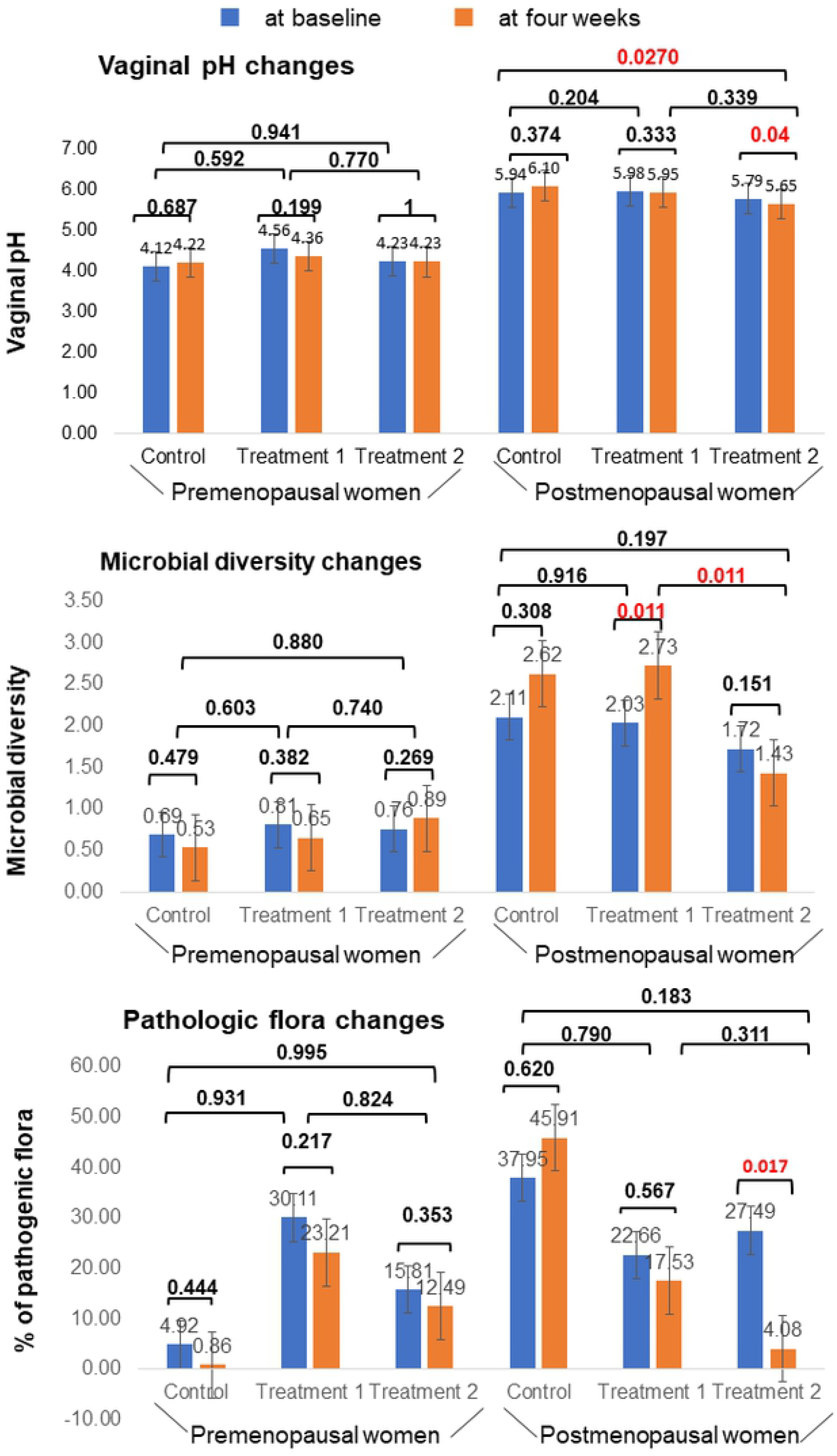
Changes in vaginal pH, microbial diversity and the proportions of vaginal pathogenic flora at baseline and four weeks after intervention. Pathogenic flora population was significantly decreased in the postmenopausal group using three feminine hygiene products.

Figure 5A showed that no one in the control group reported improvement in genitourinary symptoms, but these symptoms significantly improved in treatment 2 group of premenopausal women (0% versus 60%, p=0.043) and treatment 1 group of postmenopausal women (0% versus 81.3%, p<0.01) compared to the control group. Among

**Figure. 5.**
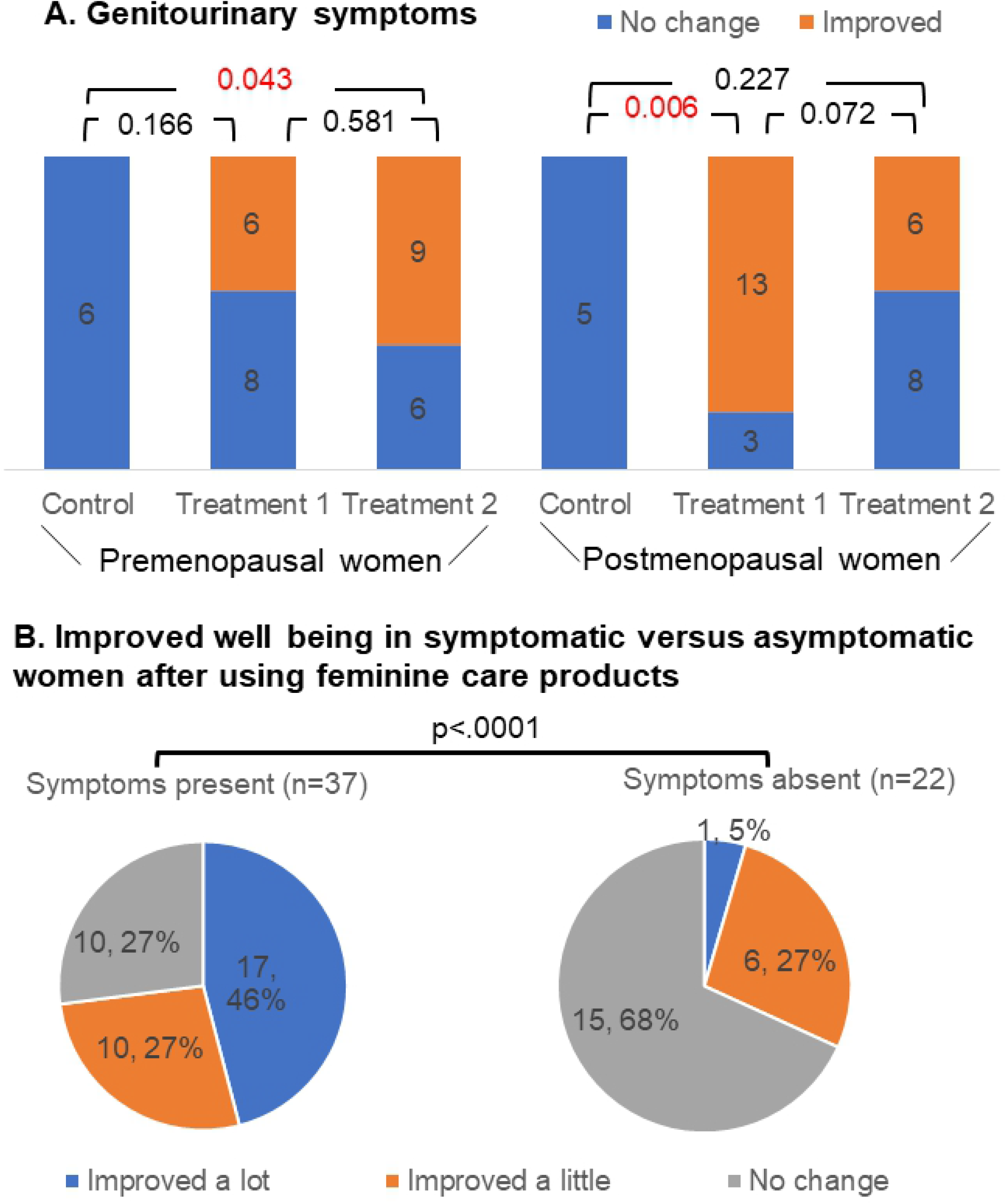
Changes in genitourinary symptoms after 4 weeks. Genitourinary symptoms significantly improved in treatment 2 group of premenopausal women and treatment 1 group of postmenopausal women compared to the control group.

37 symptomatic women, 27 (73%) reported that their symptoms got improved after undergoing either treatment 1 or treatment 2. Even among 22 asymptomatic women, 32% (n=6) reported better wellbeing. In premenopausal women, the most improved symptom reported among treatment 1 group was odor and that was discharge among treatment 2 group. In post-menopausal women, both groups reported improvement in a wide variety of symptoms.

As shown in figure 6, the average simplified menopausal index scores showed only marginal changes except in premenopausal treatment 2 group where mild menopausal symptoms got improved after 4 weeks (23 versus 15, p<0.005). The overactive bladder symptom scores did not change much before and after 4-week treatment in all groups. Among 12 postmenopausal women in the intervention groups who had baseline OABS scores of 3 and above (overactive bladder suspect), 11 women had significantly reduced OABS scores after using feminine hygiene products.

**Figure. 6.**
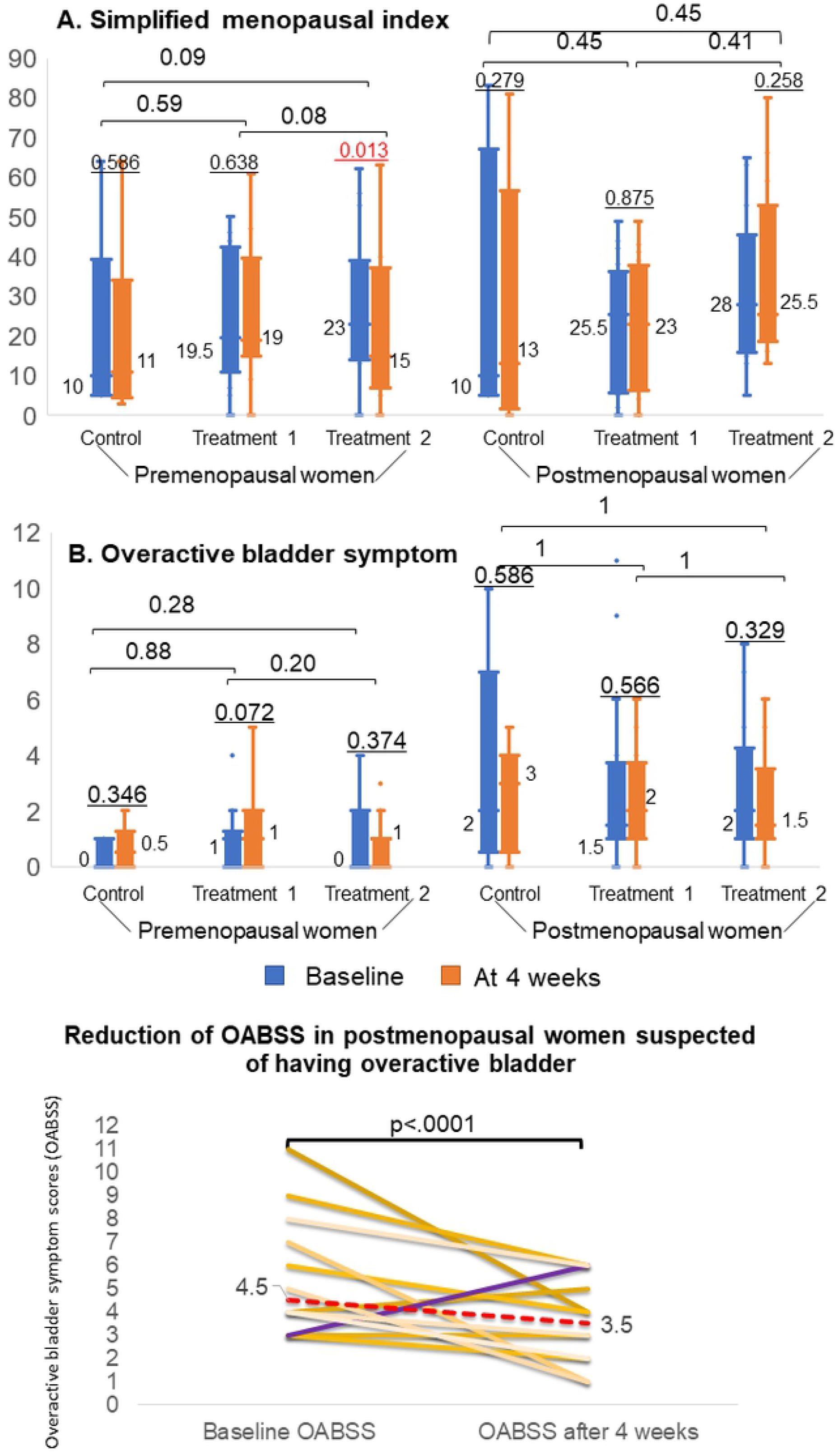
Changes in median symptom scores after 4-week treatment with feminine care products. Simplified menopausal index scores in premenopausal treatment 2 group reduced after 4 weeks. Significantly improved OABB symptom scores were seen in those with OABB symptom scores more than 3 in postmenopausal women after 4 weeks of treatment.

## Discussion

Genitourinary symptoms of menopause (GSM) are the new terms for a broad spectrum of symptoms comprising of urinary symptoms and sexual symptoms in addition to the existing vulvovaginal atrophy (VVA) symptoms. It was first introduced in 2014 by a consensus of the International Society for the Study of Women’s Sexual Health and the North American Menopause Society. More than half of the women in the middle age were reported of having GSM.^11^ Since it is a chronic or progressive condition, which cannot be healed by nature, specific treatment is usually necessary. Symptoms usually result from atrophic vaginitis due to aging changes. However, even in young women, dryness and vulva discomfort can be significant symptoms. Therefore, GSM should not be considered as the condition that affects the postmenopausal women only. Daily preventive measures should be in place as they can improve the quality of life (QOL) of a wide range of women.

In this study, approximately 60% of healthy women in each premenopausal and postmenopausal group experienced at least one genitourinary symptom. Postmenopausal women had worse symptoms and 50% of them reported that these symptoms affected their daily lives significantly. After using feminine hygiene products, genitourinary symptoms got significantly improved in the treatment 2 group of premenopausal women and treatment 1 of postmenopausal women. At baseline, postmenopausal women had more urinary symptoms such as urinary incontinence, frequencies, etc. However, those symptoms significantly improved in postmenopausal women with OABB scores more than 3 after using feminine hygiene products.. Since genitourinary symptoms were significantly associated with overactive bladder symptoms, ^29^ improved overactive bladder symptoms could be associated with improved genitourinary symptoms in the postmenopausal women.

*Lactobacillus iners* (CST III) was the most prevalent community subtype of *Lactobacillus* in the women in our study. This was consistent with the CST prevalence studies in the previous studies on Asian women.^2^ *Lactobacillus* population was 72% in the premenopausal women, whereas it was 10% in the postmenopausal women, indicating the effect of hypoestrogenic environment after menopause. The *Lactobacillus* strains used in this study, *Lactobacillus plantarum* and *Lactobacillus rhamnosus*, exhibited antagonistic activity against the tested pathogens in vivo tests. ^19, 30, 31^ In addition, some specific strains of these species, when administered orally or intravaginally, reshaped the vaginal ecosystem and protected against vaginal infections such as bacterial vaginosis and vaginal candidiasis. ^32-36^ Consistent to these findings, after feminine product use, pathogenic flora population significantly reduced in both premenopausal and postmenopausal women. In addition, *Lactobacillus* population increased in the premenopausal women. Vaginal pH and pathogenic flora population significantly decreased in the postmenopausal group using three feminine hygiene products (Treatment 2 group). Microbial diversity was decreased in the same group although it did not yield statistical significance. These results suggest better results with direct application of vaginal lubricant gel containing *Lactobacillus* in the postmenopausal women.

As with the majority of studies, our study has a few limitations. An important limitation of this study was insufficient sample size especially in the premenopausal women, leading to low power. A post hoc power analysis was conducted using R statistical software 4.1. The analysis revealed that, for an effect size of 0.617 in vaginal pH, and 0.733 in the proportions of pathogenic flora in the premenopausal women, the statistical power of the one-way ANOVA was 45.5% and 60.2% respectively for at least 5 participants in the control group, which was significant at the 5% level (two tailed). We need at least 10 participants per group to yield a statistical power of 80% or more in the premenopausal women. We might have overestimated the effect size in our small preliminary study without a control group. Not only the sample size, but also the length of the study can limit the benefits of the treatments as the pilot study lasted longer than this study (8 weeks versus 4 weeks). With few observations and longer duration, the results of the preliminary study would possess less precision and accuracy and more effect size. Moreover, due to the nature of microbial heterogeneity among the participants, we need larger and longer trials to evaluate the effectiveness of feminine care products. We also need to assess whether intermittent or continued use of feminine hygiene products is effective since the recurrence of bacterial vaginosis is common after several months. In addition, we need to identify the probiotic strains and administrative routes that could yield the best possible outcome. Finally, similar to other clinical trials, the study results have limited generalizability and applicability. Those voluntarily enrolled in the clinical trials may not represent the real-world population. However, this study provided insights to justify sample size and study duration for a future full-scale trial.

## Conclusion

Genitourinary symptoms might be improved using feminine hygiene products containing *Lactobacillus* in a wide range of women populations. Such products can also create a better balance of *Lactobacillus* and pathogenic flora population, especially in the postmenopausal women. Direct application of *Lactobacillus* inside the vagina might produce better results. However, we need larger comparative studies with different intervention periods, probiotic strains and administrative routes to evaluate the best possible outcome of using *Lactobacillus* in the maintenance of healthy vaginal ecosystem.

## Data Availability

All relevant data are within the manuscript and its Supporting Information files.

## Acknowledgments

We would like to acknowledge all the women who willingly participated in the study.

